# Comprehensive evaluation of EEG spatial sampling, head modeling, and parcellation effects on network alterations in idiopathic generalized epilepsy

**DOI:** 10.1101/2024.11.25.24317908

**Authors:** Christina Stier, Markus Loose, Carmen Loew, Marysol Segovia-Oropeza, Sangyeob Baek, Holger Lerche, Niels K. Focke

## Abstract

Idiopathic generalized epilepsy (IGE) is characterized by marked brain network alterations as assessed using electrophysiology. Logistical challenges and the need for a volumetric MRI often hinder the clinical application of high-density EEG or MEG. This study investigates the influence of EEG channel density and the head model on brain metrics derived from 256-channel EEG and 19-channel routine EEG in two samples balanced for age and sex. First, we evaluated resting-state data from 35 individuals with IGE and 54 healthy controls collected using the 256-channel setup. Data were analyzed at full density and then iteratively down-sampled to lower densities. Source reconstruction was performed either using individual MRI data or a standard brain template and dynamic imaging of coherent sources (DICS). We assessed EEG power and connectivity (imaginary part of coherency) group differences at all channel compositions, head model types, and parcellations (cortical vertices, anatomical and network parcellations). Second, a routine sample recorded with 19 channels was analyzed to validate findings in a real epilepsy monitoring scenario (71 patients, 43 controls). We found that lower-density arrays reliably identified global group differences for both power and connectivity and in frequency bands for which the strongest effects were observed. The spatial similarity of the results for the 256 channels set and those with fewer channels were good to moderate for power (r_spin_ ∼0.97 to 0.33), but dropped for connectivity with fewer than 64 channels (r_spin_ ∼0.78 to-0.12). Comparing individual and canonical head models revealed consistent effects (r_spin_ ∼0.77 to 0.5), with coarser brain parcellations increasing stability for low-density maps. In sum, low-density EEG arrays suffice for detecting global alterations in IGE, particularly in signal power. Our findings advocate for leveraging clinical EEG for brain-wide analyses in IGE while emphasizing the need for high-density coverage if spatial precision is needed. Canonical head models are a viable alternative if no individual MRI is available, especially for regional-or network-level assessments.

## 1. Introduction

Since the invention of EEG, the EEG systems have undergone a constant development and remained an essential tool in the diagnostics of epilepsy. Today, systems with a dense spatial coverage are available and generally expected to lead to a better spatial resolution than sparse arrays. The current guidelines by the International Federation of Clinical Neurophysiology (IFCN) in 2017 have been tailored for the clinical diagnostic use, which recommends the use of at least 25 electrodes in a standard array, and the 10-10 system or high-definition systems with 64 to 128 electrodes for source localization purposes (Seeck et al., 2017). Several studies have demonstrated improvements in the source localization of interictal or ictal epileptic discharges using high-density recordings (Brodbeck et al., 2011, Lantz et al., 2003, Song et al., 2015). In other (single) cases, the localization of seizure onset zones (Staljanssens et al., 2017) and the mapping of the language areas improved (Stoyell et al., 2021), and epileptic oscillations at rapid scales could be identified (Stoyell et al., 2021). Others argue that high-density EEG (HD-EEG) not necessarily improves diagnostic accuracy if the number of spikes is sufficiently high (Vorderwülbecke et al., 2021) or data recordings long enough (Justesen et al., 2019).

In general, evaluations of high-density over low-density systems have been limited to diagnostics in the presurgical settings and patients with suspected focal epilepsies. However, in recent years, a network perspective of epilepsy and its mechanisms has emerged and prompted a series of studies investigating whole-brain networks in generalized and focal seizure types. In this context, the effects of spatial sampling on brain-wide mapping of source-reconstructed EEG signals have not been sufficiently examined. While there is accumulating evidence for altered interictal, spectral network alterations in patients with IGE (Elshahabi et al., 2015, Li Hegner et al., 2018, Niso et al., 2015, Stier et al., 2022), it is unclear to what extent these results can be expected for data derived from a clinical setting. The clinical standard in epilepsy monitoring units is often still limited to the conventional low-density recordings as it is less costly and time-intensive than a high-density montage. To leverage the wealth of clinical EEG databases and assess the potential of resting-state brain dynamics for diagnostic purposes, a side-by-side evaluation of data acquired in clinical and research settings using similar methods is essential. Advanced brain-wide modelling should also provide spatial insights into the underlying effects, enabling differentiation between epilepsy syndromes and other neurological disorders. Furthermore, in resource-restricted countries and individuals with IGE, volumetric MRI scans may not be available for precise source reconstruction.

We therefore set out a study to compare resting-state brain activity and synchronization between patients with IGE and controls measured using a 256-channel EEG. We focused on two well-established metrics such as power (Clemens et al., 2000, Elshahabi et al., 2015, Miyauchi et al., 1991, Pegg et al., 2020) reflecting the squared signal amplitude, and the imaginary part of coherency (Nolte et al., 2004) as our connectivity measure, since liability to pathological synchronization has been described even during the interictal state (Segovia-Oropeza et al., 2025, Stier et al., 2021, Tangwiriyasakul et al., 2019, Tangwiriyasakul et al., 2018). We subsequently repeated the group-level analyses after virtually reducing the number of channels, applying conventional source reconstruction based either on individual MRI anatomy or on a brain template (hereinafter referred to as the “canonical” head model). Although accurate tissue boundaries generally improve source reconstruction solutions (Henson et al., 2009, Huiskamp et al., 1999) and can enhance localization in focal epilepsy (Brodbeck et al., 2011), the added value for whole-brain, group-level studies in generalized epilepsies remains unclear. To mirror previous work in IGE, we first tested group differences at brain surface vertices and then repeated the analyses after aggregating data into anatomical regions and functional networks, evaluating whether coarser parcellations yield more consistent results across channel densities. Finally, we validated the findings in an independent cohort recorded with a routine 19-channel EEG, thereby assessing the effects under typical clinical conditions. The overarching aim of this work is to provide an overview of spatial sampling effects on brain metrics commonly used to quantify neurophysiological alterations in epilepsy.

## 2. Materials and Methods

### 2.1. Participants

We considered healthy individuals and individuals diagnosed with IGE according to the International League Against Epilepsy (Scheffer et al., 2017) from two sites. Participants included in the HD-EEG sample were recruited and measured through the Department of Neurology, University Hospital of Tübingen, Germany, between 2013 and 2019, or through the Clinic of Clinical Neurophysiology, University Medical Center of Göttingen, Germany, between 2018 and 2020. The local ethics committee of the Medical Faculties in Tübingen (ethics number 646/2011BO1) and Göttingen (ethics number 16/10/17) approved the studies, compliant with the Declaration of Helsinki. In total, 35 patients and 54 controls were included in the HD-EEG sample, of which data from 22 patients and 34 controls were re-analyzed from a previous study (Stier et al., 2022). The second sample included 71 patients and 43 controls undergoing a routine clinical procedure (see section 2.2.). These data were acquired between 2007 and 2021 in the Clinic of Clinical Neurophysiology, Göttingen. The need for informed consent was waived for this retrospective study by the local ethics committee (ethics number 2/5/21). In general, only participants with normal MRI scans or nonspecific findings (e.g., cysts) were considered in the study. Clinical information can be found in Table 1. All controls were free of neurological or psychiatric conditions, and none of them were taking any medication at the time of the measurement. The two samples were comparable in age (F_HD-EEG_ = 0.17, p_HD-EEG_ = 0.68; F_routine_ = 0.92, p_routine_ = 0.34) and sex (χ^2^_HD-EEG_= 0.0, p_HD-EEG_= 1; χ^2^_routine_= 0.93, p_routine_= 0.33).

**Table 1.** Demographics and clinical information.

|  | Main study sample |  | (Clinical) routine sample |  |
| --- | --- | --- | --- | --- |
|  | Patients | Controls | Patients | Controls |
| Total (n) | 35 | 54 | 71 | 43 |
| Site: total n Göttingen/Tübingen | 10/25 | 15/39 | 71/0 | 43/0 |
| Female, n (%) | 18 (72) | 27 (49) | 35 (49) | 26 (60) |
| Mean age (y), (sd) | 32 (11.6) | 31 (12.2) | 29 (10.2) | 28 (8.8) |
| GSWD during EEG recordings, n (%) | 13 (37) | 0 (0) | 19 (27)<sup>a</sup> | 0 (0) |
| Seizure free > 12 months, n (%) | 19 (54) | - | 55 (77) | - |
| Drug intake at measurement, n (%) | 2 (6) | - | 13 (10) | - |
| Epilepsy syndrome, n (%) |  |  |  |  |
| CAE | 5 (14) | - | 3 (4) | - |
| JAE | 10 (29) | - | 7 (10) | - |
| JME | 7 (20) | - | 23 (32) | - |
| GTCS | 6 (17) | - | 19 (27) | - |
| IGE | 7 (20) | - | 19 (27) | - |

Abbreviations: CAE = childhood absence epilepsy; GSWD = generalized spike-wave discharges; GTCS = generalized tonic clonic seizures only; IGE = genetic generalized epilepsy (unclassified); IQR = interquartile range; JAE = juvenile absence epilepsy; JME = juvenile myoclonic epilepsy; sd = standard deviation; y = years. <sup>a</sup> Including eight patients who showed hyperventilation-induced GSWD.

### 2.2. EEG recordings

Individuals from the HD-EEG sample underwent 30 minutes of continuous resting-state recordings using a 256-channel system (GES400; Magstim EGI) with a sampling frequency of 1 kHz. All individuals were instructed to keep their eyes closed while relaxing, not to fall asleep, and not to think of anything in particular. For the routine sample, a 19-channel EEG with classical 10-20 montage (Jasper, 1958, Klem, 1999) was used with sampling frequency of 500 Hz and 20 minutes of recording time conducted according to clinical standard in the routine neurophysiological laboratory. As such, the patients of this sample underwent a routine clinical procedure with periods of hyperventilation, tests for the Berger effect, and in a few cases intermittent photic stimulation. Controls for the clinical sample were recorded in the same laboratory with the same equipment, and tests for Berger effect were equally applied, but no hyperventilation and photic stimulation.

### 2.3. Head models

In the HD-EEG study, each individual had a sagittal T1-weighted MRI acquisition (3D-MPRAGE, repetition time = 2.3 s, echo time = 3.03 ms, flip angle = 8° (Tübingen) or 9° (Göttingen), voxel size = 1 × 1 × 1 mm) subsequent to the EEG recordings either using a 3T Siemens Trio (12-channel headcoil; 12/54 controls; 4/35 patients) or Prisma/Prisma^fit^ scanner (64-channel headcoil; 42/54 controls; 31/35 patients). As anatomical information was not available for all individuals of the routine sample, we used an MRI template for all individuals based on 225 T1 and FLAIR images with a large field-of-view customized for EEG head model generation (Kreilkamp et al., 2023). To project sensor level data onto cortical surfaces, we applied FreeSurfer reconstruction to T1 images and used SUMA (Saad and Reynolds, 2012) to resample the cortical surfaces (density factor = 10) based on a standard template (‘fsaverage’). This procedure allowed anatomical correspondence between individuals at 1,002 vertices per hemisphere serving as our source points. Volume conduction models were built using the individual or template-derived cortical meshes, brain segmentations via SPM12 processing (Ashburner and Friston, 2005), and the EEG electrodes realigned to the anatomical landmarks (nasion, preauricular points) and projected onto the scalp mesh. We used a three-layer boundary element model (‘openmeeg’ (Gramfort et al., 2010)) to compute the leadfields using Fieldtrip (Oostenveld et al., 2011) in MATLAB (R2018b).

### 2.4. EEG processing and source analysis

Using Fieldtrip (Oostenveld et al., 2011), we filtered the raw data (first-order Butterworth filter at 1 and 70 Hz, line-noise removal at 50 Hz and its harmonics), downsampled to 150 Hz, demeaned, and segmented the data into trials of 10 s length. All EEGs were re-referenced to a common average reference online. Trials were visually inspected and those containing artefacts (movements, muscle artefacts, sensor jumps) rejected. In case of spike-wave discharges occurring during the EEG recordings, the respective, the preceding and following trial were removed. In the routine EEG sample, only trials with eyes-closed and trials without external stimulation parameters plus 90 seconds recovery were considered for further analysis. We applied independent component analyses to detect and (manually) reject components with cardiac and eye movements. All remaining trials were reviewed a second time and vigilance of the individuals was rated according to the criteria of the American Academy of Sleep Medicine (https://aasm.org/). Thirty clean data trials rated as awake were then randomly drawn for each individual as we have shown that EEG power and connectivity metrics are reliable for five minutes of data (Marquetand et al., 2019). To project the data from the electrode-level to the surface points, we applied fast Fourier spectral analyses using multitapers (DPSS) for six frequency bands (center frequency ± smoothing: delta: 2 ± 2 Hz, theta: 6 ± 2 Hz, alpha 10 ± 2 Hz, beta1 16 ± 4 Hz, beta2 25 ± 4 Hz and low gamma 40 ± 8 Hz). Based on this, cross-spectral densities and EEG power were estimated and frequency-dependent source projection performed using Dynamic Imaging of Coherent Sources (Gross et al., 2001) (regularization: 5%). At each frequency band, coherency between all pairs of vertices (n = 2004) and power was computed for each vertex. The absolute imaginary part of the coherency coefficient was used as connectivity measure in this study to account for potential fieldspread (Nolte et al., 2004). All connections between vertices were then averaged to yield an estimate of synchronization of a vertex, and, for individual global estimates, also across all vertices.

### 2.5. Channel reduction and statistical procedure

To test the impact of spatial sampling on statistical comparisons between the patients and controls, electrode layouts were created with 192-, 128-, 64-, 32-channel maps, and maps with 19 and 25 channels based on the original 10-20 (Jasper, 1958, Klem, 1999) and extended 10-20 montage (Seeck et al., 2017), respectively, following the guidelines of the IFCN. The full 256-channel layout was used as original template. For the 192 to 48 channel-maps, channels from the original 256-layout were removed iteratively based on the distance to neighboring channels. Corresponding channels positioned on the other hemisphere were equally removed. Channels assigned to the extended 10-20 positions were kept in the reduction process (see Figure 1). For each channel set, the cleaned electrode-level data of each individual in the HD-EEG study sample was selected, leadfields re-computed and source projection repeated.

**Figure 1.**
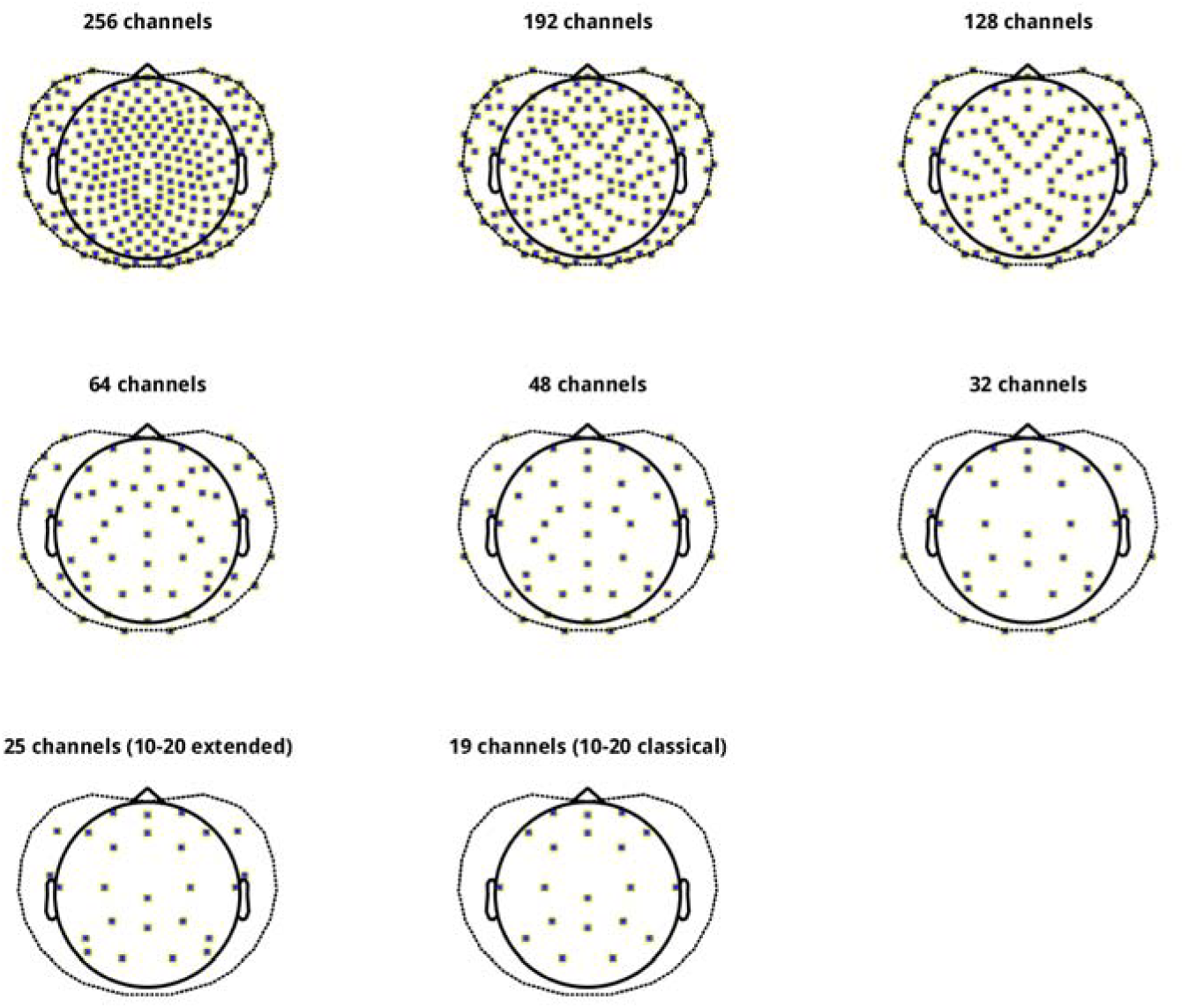
Channel compositions at different densities Shown are the channel layouts used to test the impact of spatial sampling on group comparisons in the HD-EEG study sample. All compositions rely on the original EGI Magstim 256-layout as a template. The number of channels was reduced in an iterative process, yielding compositions of different densities. Channels assigned to the positions of the classical 10-20 montage were kept throughout (see Methods in the main manuscript for further details). The outline of the layout was provided by the Fieldtrip toolbox (Oostenveld et al., 2011).

### 2.6. Group-level statistics

To assess whether patients differed from controls in power and connectivity, we tested one-sided contrasts using permutation analyses of linear models using PALM (Winkler et al., 2014) at each of the surface-vertices and once for the global metrics. Age, sex, and scanner site were included as regressors of no interest. P-values were computed from the resulting empirical distribution and threshold-free cluster enhancement (TFCE) (Smith and Nichols, 2009), and familywise error (FWE) corrected at the cluster level. We used tail approximation with 5000 permutations for accelerated inference (Winkler et al., 2016). The significance level was set to –log_10_(p) = 1.3 (equivalent to p <.05). This statistical procedure was carried out for each channel set in the HD-EEG sample as well as for the routine sample measured using low-density EEG. To determine standardized group mean differences, we used Cohen d computed on the t-values of the group factor of the linear models adjusted for the effects of the covariables. A d ≥ 0.8 indicates a large, d = 0.5 a medium, and d = 0.2 a small effect.

### 2.7. Spatial correlations of effect size maps

To assess the similarity of the group-level results among the different channel sets, we correlated the original effect size map (256 channels) with the maps for the reduced channel sets (192 to 19 channels). We focused on the EEG differences in the theta band, as this has yielded the strongest and most consistent results in the present study and others (Faiman et al., 2021). We applied the spin-test (Alexander-Bloch et al., 2018) to assess statistical significance of the Spearman rank coefficients based on 1,000 random rotations of the spherical cortical surface of each hemisphere separately as it takes the spatial embeddedness of the correlated cortical maps into account.

### 2.8. Channel densities across parcellation schemes

To evaluate group differences (IGE vs. controls) at different cortical resolutions, we remapped individual power and connectivity by averaging across vertices within each of 68 brain regions defined by Desikan et al. (2006) and 14 functional resting-state networks defined by Yeo et al. (2011). We then ran separate analyses for each parcellation scheme using PALM (see section 2.6.) and, as described above (section 2.7.), computed spatial correlations between the results for the 256-channel maps and the maps for the virtually reduced channel sets (192 to 19 channels). To ensure comparability of the results among the different resolutions, we standardized the correlation coefficients using Fisher’s z transformation (Fisher, 1921) and estimated the probability of the z-differences between the vertex-space and the Desikan regions and Yeo networks, respectively, using R (Team, 2013). We controlled the false discovery rate (FDR) for seven comparisons (original versus lower-density maps) at each type of head model and EEG metric.

### 2.9. Impact of the head model type

We further tested whether the group-level results for the HD-EEG sample were different depending on whether individual or canonical head models were used. For this purpose, the respective Cohen d maps for the results in the theta band were correlated within each channel set using the spin-test as described in 2.7. (1,000 permutations). Again, Fisher’s z transformation of the coefficients and FDR correction of the p-values were applied at the level of channel sets (n = 8) to compare the effects across parcellation schemes (vertices, regions, networks).

## 3. Results

### 3.1. Group contrast at vertex-based resolution (individual head models)

Group contrasting at the vertex resolution using a 256-channel EEG and individual head models yielded significantly higher connectivity and power in patients with IGE than in controls. For connectivity, this was mainly the case in the theta frequency band in frontal, centrotemporal, and posterior brain regions (Figure 2A) and also globally (d_theta_ = 0.76, p_theta_ < 0.001), indicating overall increased levels in the patients. Weaker patterns were also observed in the delta and alpha frequency bands (Figure 2A), which reached significance at the global level only in delta (d_delta_ = 0.62, p_delta_ < 0.01). There were no vertex-wise or global significant differences in the beta and gamma bands (p > 0.05). Increased power in the patients was significant across the brain and the frequency spectrum (delta to gamma, global d: 0.83-1.12, p < 0.001), with emphasis on posterior brain regions (Figure 2B).

**Figure 2.**
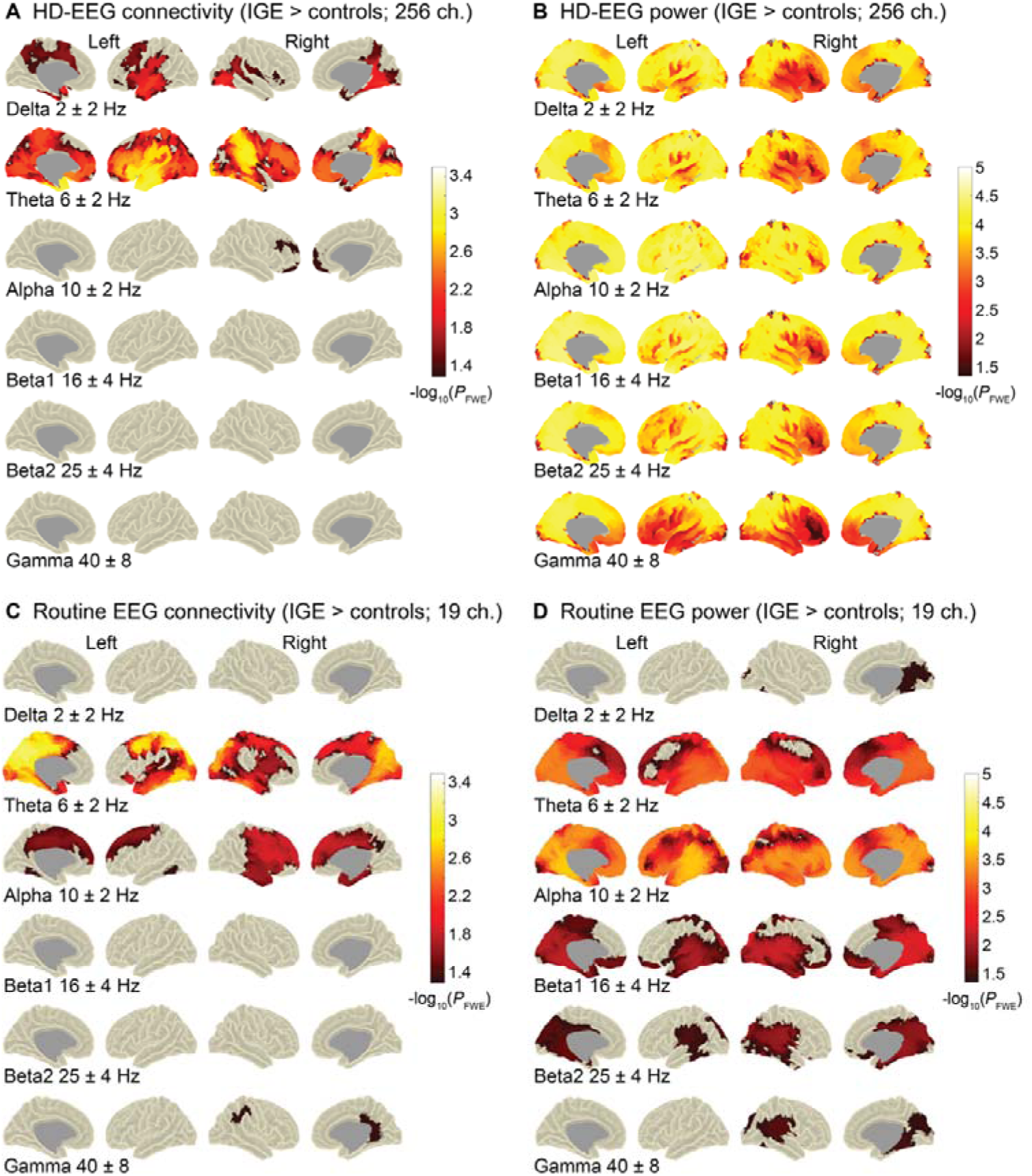
Vertex-based group comparisons using high-density (256 channels) and low-density EEG (19 channels) Highlighted are vertices on the cortical surface for which the patients with IGE from the HD-EEG sample (n = 35) and the routine sample (n = 71), respectively, had significantly higher connectivity (A, C) and power (B, D) than the controls (n_HD-EEG_ = 54, n_routine_ = 43). Age, sex, and measurement site were included as covariates into the permutation-based and frequency-specific analysis of linear models. The significance level was set at-log10(p) = 1.3 (equivalent to p = 0.05). Shown are the results after familywise error correction (FWE) and threshold-free cluster enhancement (TFCE). For the HD-EEG sample (A, B), the analyses were performed using individual head models and 256 EEG channels. The routine sample (C, D) was measured using 19 channels with a classical 10-20 montage and analyses were performed using a template head model as described in the Methods. Similar to the results for the HD-EEG sample, the strongest effects were observable for theta connectivity. In the same vein, increased power in the patients was distributed across the frequency spectrum with a posterior focus, but with generally weaker effects than in the HD-EEG study sample. Ch. = channels; center frequency ± smoothing using multitapers.

### 3.2. Differences in global levels among channel sets

To provide a global estimate and comparison of the contrast IGE > controls for different channel sets (Figure 1), we report standardized group mean differences in Figure 3 (Cohen d). For global connectivity, the significance threshold (p_FDR_ < 0.05) was exceeded for all channel densities only in the theta band (d > 0.5), irrespective of the head model type used. Increased connectivity in the delta frequency band was observed for 256-and 192-channels using an individual head model (d > 0.5, p_FDR_ < 0.05) and also in the beta1 band for both head models (individual: d > 0.4 for 192-48 channels; canonical: d > 0.4 for 256-25 channels; p_FDR_ < 0.05). No significant differences in any of the channel sets and head model types were observed in the remaining frequency bands, i.e., alpha, beta2 and gamma (d < 0.3, p_FDR_ > 0.05). Increased global power in the patients was significant for all channel sets and head models (individual: d > 0.8; canonical: d > 0.7).

**Figure 3.**
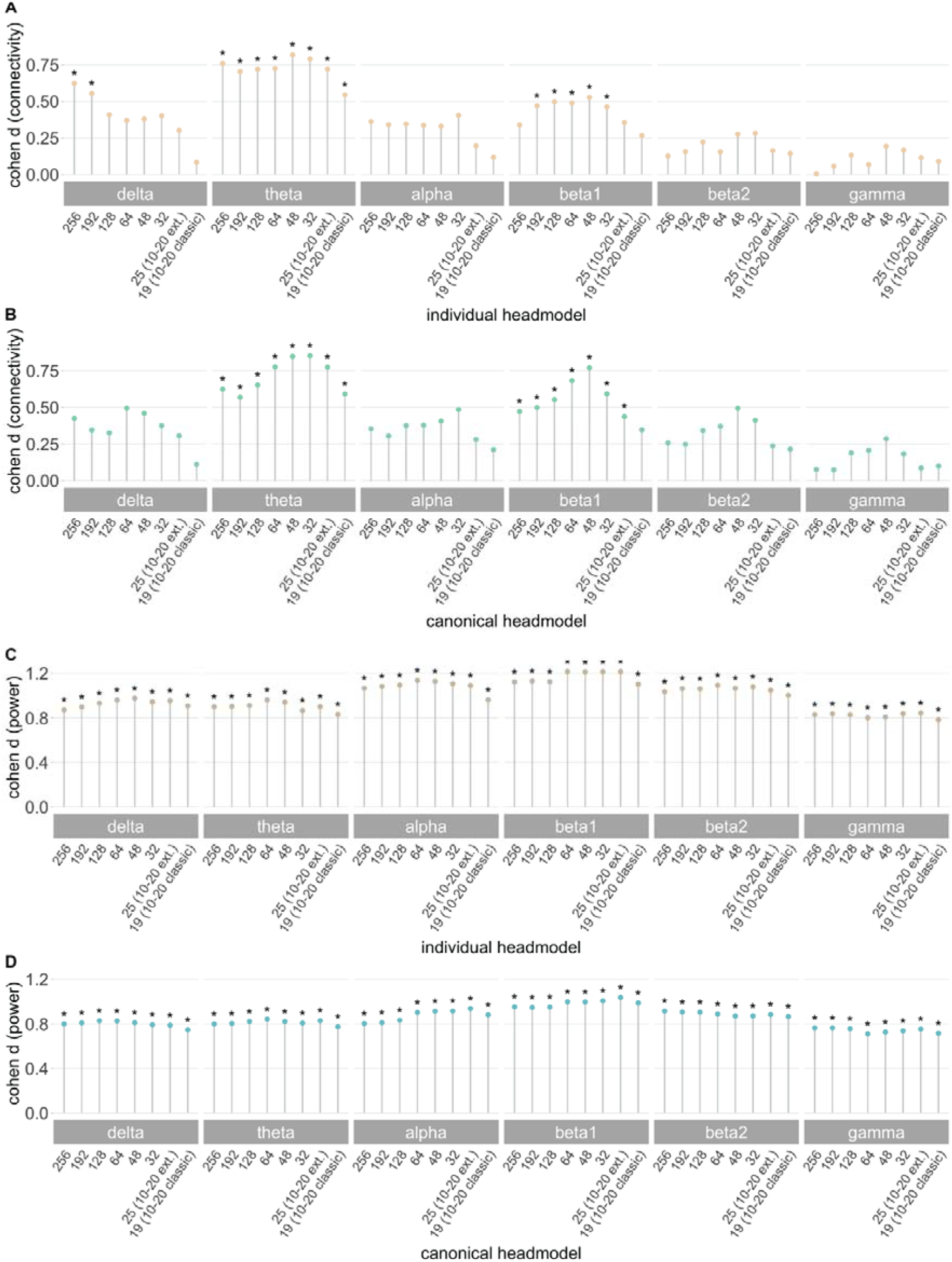
Group differences (IGE > controls) in global connectivity and power at different channel densities Each colored dot represents standardized effect sizes (Cohen d) for increased connectivity (A, B) and power (C, D) in the patients with IGE (n = 35) compared with the controls (n = 54) of the HD-EEG sample based on permutation analysis of linear models. Separate analyses were run for each channel density and head model type (beige: individual, turquoise: canonical). All d-values were computed based on the respective t-value of the group factor and corrected for the influence of age, sex, and measurement site. A d ≥ 0.8 indicates a large, d = 0.5 a medium, and d = 0.2 a small effect and allows direct comparison of the results for EEG metrics and head model types. Statistically significant group comparisons after correction for multiple comparisons at the level of channel sets within each frequency band are indicated with an asterisk (*p_FDR_ <.05). Electrode layouts were created with 192-, 128-, 64-, 32-channel maps, and maps with 19 and 25 channels based on the original 10-20 (10-20 classic) and extended 10-20 montage (10-20 ext.).

### 3.3. Effect of spatial sampling for vertex-connectivity (theta band, individual head models)

To quantify to what extent the spatial EEG patterns vary depending on the channel density, we focused on the group contrasts in the theta frequency band (IGE > controls) as this turned out to be the strongest global effect across channel sets and head model types. The virtual channel reduction generally influenced the group contrast in source space stronger for connectivity than for power (Figure 4). Correlations between connectivity maps (Cohen d) for 256 channels and those from 192-and 128-channels were high (r_s_ = 0.77-0.43, p_spin_fdr_ < 0.05), but dropped to r_s_ = 0.21-0.09 (p_spin_fdr_ > 0.05) with 64 channels and fewer (Figure 4). To visualize spatial differences, we subtracted the effect size maps for each channel density from that of 256 channels (Figure S1). The high-density sampling revealed stronger connectivity differences in parietal and frontal brain areas compared to lower channel densities. Conversely, the effect sizes for fronto-central and temporal regions were larger at low channel densities than at high densities. In general, the sampling strategies differed at a maximum of d ± 0.6 from the full set.

**Figure 4.**
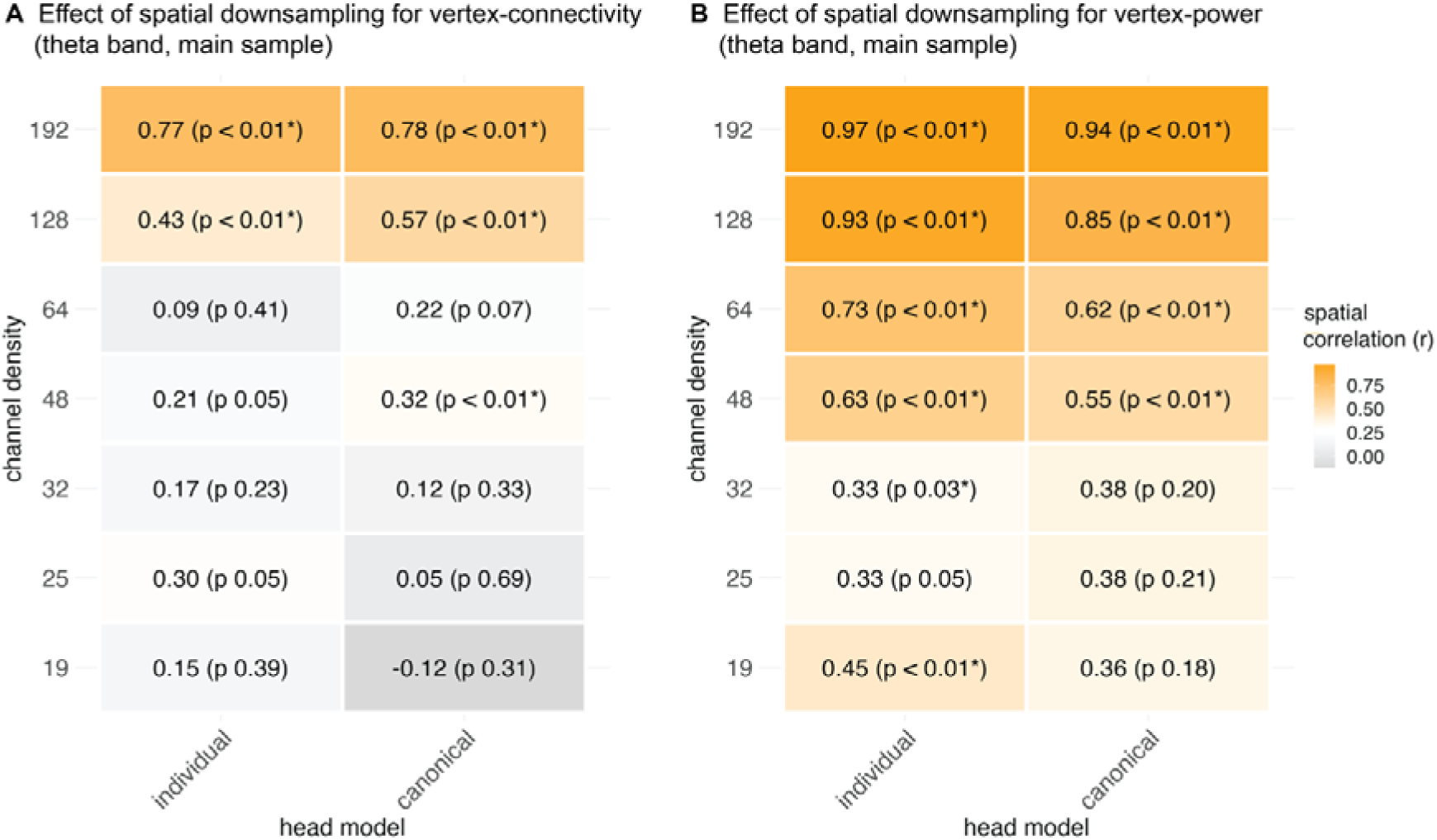
Spatial correlation analyses of group contrasts in the theta band across channel densities We used Spearman’s rank correlation (r) to quantify the spatial similarity between the effect size maps (Cohen d) for EEG levels (IGE > controls) based on the 256-channel density and each lower-density channel set (192-19 channels). The correlation analyses were performed for the group contrasts at 2004 cortical vertices. * Indicates significance of the correlations for the vertex-space using the spin-test and after controlling the FDR at the level of channel set comparisons (n =7). P_spin_-values are indicated in brackets.

### 3.4. Effect of spatial sampling for vertex-power (theta band, individual head models)

For comparability with the connectivity results, we focus on the theta frequency band for power as well. The correlations between the effect size maps at 256 channels for theta power and the maps at the other channel sets were significant for all densities except for 25 channels (r_s_ = ∼0.97-0.45, p_spin_fdr_ < 0.05; r_s_25-channels_ = 0.33, n.s., Figure 4). In terms of spatial variability, full sampling (256 channels) yielded stronger power differences in the prefrontal cortex than for the lower densities, particularly for 48 channels and below, but smaller differences in the middle temporal areas (Figure S2).

### 3.5. Sampling effects across parcellation schemes

We further tested whether remapping of EEG metrics from the vertices to anatomical regions or functional networks were more robust to the virtual reduction of channels. Thus, we statistically compared the transformed correlation coefficients derived from the comparisons between the d-maps at 256 channels and the other channel sets (Table 2). The correlations for connectivity did not differ between the vertex-resolution and the Desikan-Killiany parcellation and the Yeo network-resolution, respectively (p_fdr_ > 0.05, individual and canonical head models). The regional resolution yielded significantly higher coefficients than at the vertices for power at the 192-(individual and canonical head model) and 128-channel densities (canonical head model). Similarly, higher coefficients were observed for the network-than for the vertex-resolution for the 192-to 64-channel sets (canonical head models), suggesting that the group results for power at the high-density sets were more stable using anatomically defined parcellations.

**Table 2.** Results for different parcellations: spatial correlation analyses of group contrasts in the theta band across channel densities.

|  | Channels | Individual head model |  |  | Canonical head model |  |  |
| --- | --- | --- | --- | --- | --- | --- | --- |
|  |  | z<br>Vertices | z<br>Regions<br>(Desikan<br>-Killiany) | z<br>Networks<br>(Yeo) | z<br>Vertices | z<br>Regions<br>(Desikan<br>-Killiany) | z<br>Networks<br>(Yeo) |
| Connectivity<br>(theta band) | 192 | 1.02* | 1.29 | 0.74 | 1.05* | 1.22 | 1.07 |
|  | 128 | 0.46* | 0.47 | 0.18 | 0.65* | 0.65 | 0.76 |
|  | 64 | 0.09 | 0.20 | 0.03 | 0.22 | 0.20 | 0.21 |
|  | 48 | 0.21 | 0.19 | -0.12 | 0.33* | 0.31 | 0.01 |
|  | 32 | 0.17 | 0.37 | 0.33 | 0.12 | 0.23 | 0.12 |
|  | 25 | 0.31 | 0.22 | 0.20 | 0.05 | 0.17 | 0.04 |
|  | 19 | 0.15 | 0.18 | -0.09 | -0.12 | 0.09 | -0.34 |
| Power<br>(theta band) | 192 | 2.09* | 2.65<sup>a</sup> | 2.65 | 1.74* | 2.09<sup>a</sup> | 2.65<sup>a</sup> |
|  | 128 | 1.66* | 1.83 | 1.95 | 1.26* | 1.66<sup>a</sup> | 2.30<sup>a</sup> |
|  | 64 | 0.93* | 1.02 | 1.53 | 0.73* | 0.95 | 1.74<sup>a</sup> |
|  | 48 | 0.74* | 0.78 | 1.07 | 0.62* | 0.73 | 1.33<sup>a</sup> |
|  | 32 | 0.34* | 0.18 | 0.02 | 0.40 | 0.41 | 0.41 |
|  | 25 | 0.34 | 0.22 | -0.01 | 0.40 | 0.40 | 0.37 |
|  | 19 | 0.48* | 0.40 | 0.33 | 0.38 | 0.38 | 0.48 |

We used Spearman's rank correlation ( $r_s$ ) to quantify the spatial similarity between the effect size maps (Cohen d) for EEG levels (IGE > controls) based on the 256-channel density and each lower-density channel set (192-19 channels). The correlation analyses were performed separately for each parcellation scheme, that is, for the group contrasts at 2004 cortical vertices, at 72 brain regions (Desikan et al., 2006), and 14 functional resting-state networks (Yeo et al., 2011). The correlation coefficients were standardized (z) using Fisher's z transformation for comparability between the parcellations. \* Indicates significance of the correlations for the vertex-space using the spin-test and after controlling the FDR at the level of channel set comparisons ( $n = 7$ ). <sup>a</sup> indicates if the transformed correlation coefficients for the regional and network analyses, respectively, significantly differed from those in the vertex-space after FDR corrections. The significance threshold was set at $p_{\text{FDR}} < 0.05$ .

### 3.6. Comparison of head model types

We assessed the influence of the head model (individual versus canonical) on the group-level results in the theta band (Table 3). In the vertex-space, the effect size maps based on individual head models significantly correlated with those based on the canonical head model for each channel set. This was the case for EEG connectivity (r_s_ = 0.50-0.73, p_spin_fdr_ < 0.05) and power (r_s_ = 0.63-0.77, p_spin_fdr_ < 0.05). The regional resolution yielded higher concordance between the types of head model than the vertex-resolution for connectivity (192-64 and 25 channels) and power (all channel sets). The network-based (Yeo) analysis also provided higher correlation coefficients than in the vertex-space, but only for power in a few channel sets (256-48 and 19 channels).

**Table 3.** Spatial correlation of the head model types across channel densities and parcellations.

| | Channel sets<br>(Individual ~<br>canonical head<br>model maps) | $r_s$<br>Vertices | $p_{spin}$<br>Vertices | $z$<br>Vertices | $z$<br>Regions<br>(Desikan-Killiany) | $z$<br>Networks<br>(Yeo) |
| --- | --- | --- | --- | --- | --- | --- |
| Connectivity<br>(theta band) | 256 | 0.50 | < 0.01* | 0.55 | 0.81 | 1.00 |
|  | 192 | 0.60 | < 0.01* | 0.69 | 1.07<sup>a</sup> | 1.38 |
|  | 128 | 0.62 | < 0.01* | 0.73 | 1.02<sup>a</sup> | 0.33 |
|  | 64 | 0.57 | < 0.01* | 0.65 | 1.00<sup>a</sup> | 0.62 |
|  | 48 | 0.52 | < 0.01* | 0.58 | 0.76 | 0.00 |
|  | 32 | 0.60 | < 0.01* | 0.69 | 0.93 | 0.81 |
|  | 25 | 0.73 | < 0.01* | 0.93 | 1.22<sup>a</sup> | 1.07 |
|  | 19 | 0.65 | < 0.01* | 0.78 | 1.00 | 1.02 |
| Power<br>(theta band) | 256 | 0.63 | < 0.01* | 0.75 | 1.21<sup>a</sup> | 1.62<sup>a</sup> |
|  | 192 | 0.66 | < 0.01* | 0.79 | 1.22<sup>a</sup> | 1.42<sup>a</sup> |
|  | 128 | 0.65 | < 0.01* | 0.77 | 1.11<sup>a</sup> | 1.49<sup>a</sup> |
|  | 64 | 0.68 | < 0.01* | 0.84 | 1.38<sup>a</sup> | 1.95<sup>a</sup> |
|  | 48 | 0.77 | < 0.01* | 1.02 | 1.80<sup>a</sup> | 2.16<sup>a</sup> |
|  | 32 | 0.68 | < 0.01* | 0.82 | 1.08<sup>a</sup> | 1.01 |
|  | 25 | 0.70 | < 0.01* | 0.87 | 1.33<sup>a</sup> | 1.26 |
|  | 19 | 0.73 | < 0.01* | 0.94 | 1.33<sup>a</sup> | 1.62<sup>a</sup> |

We used Spearman's rank correlation ( $r_s$ ) to quantify the spatial similarity between the effect size maps (Cohen d; IGE > controls) estimated based on either individual or canonical head models. The correlation analyses were performed separately for each parcellation scheme, that is, for the group contrasts at 2004 cortical vertices, at 72 brain regions (Desikan et al., 2006), and 14 functional resting-state networks (Yeo et al., 2011). The correlation coefficients were standardized ( $z$ ) using Fisher's $z$ transformation for comparability between the parcellations. \* indicates significance of the correlations for the vertex-space using the spin-test and after controlling the FDR at the level of channel sets ( $n=8$ ). <sup>a</sup> indicates if the transformed correlation coefficients for the regional and network analyses, respectively, significantly differed from those in the vertex-space after FDR corrections. The significance threshold was set at $p_{FDR} < 0.05$ .

### 3.7. Validation in a clinical context

To validate the results derived from the virtual channel reduction approach, we conducted the same processing and analysis principles on data acquired in a clinical setting using a classical routine 10-20 EEG system and a canonical head model. As with the HD-EEG sample and the virtual 19-channel set, increased connectivity was observed in patients compared with controls with the most pronounced effects in the theta band and effects in alpha (Figures 2 and 5). In a few clusters, there was significantly increased connectivity in the gamma band, which was not the case in the HD-EEG analysis nor the virtual 19-channel set. Conversely, no effects were found for delta connectivity, as was the case for the HD-EEG analysis (compare Figure 2A). The power analysis of the routine EEG data yielded similar results to those obtained using the 256-channel and 19-channel sets (compare Figure 2B), except for the delta band, for which increased power patterns did not survive corrections for multiple comparisons.

**Figure 5.**
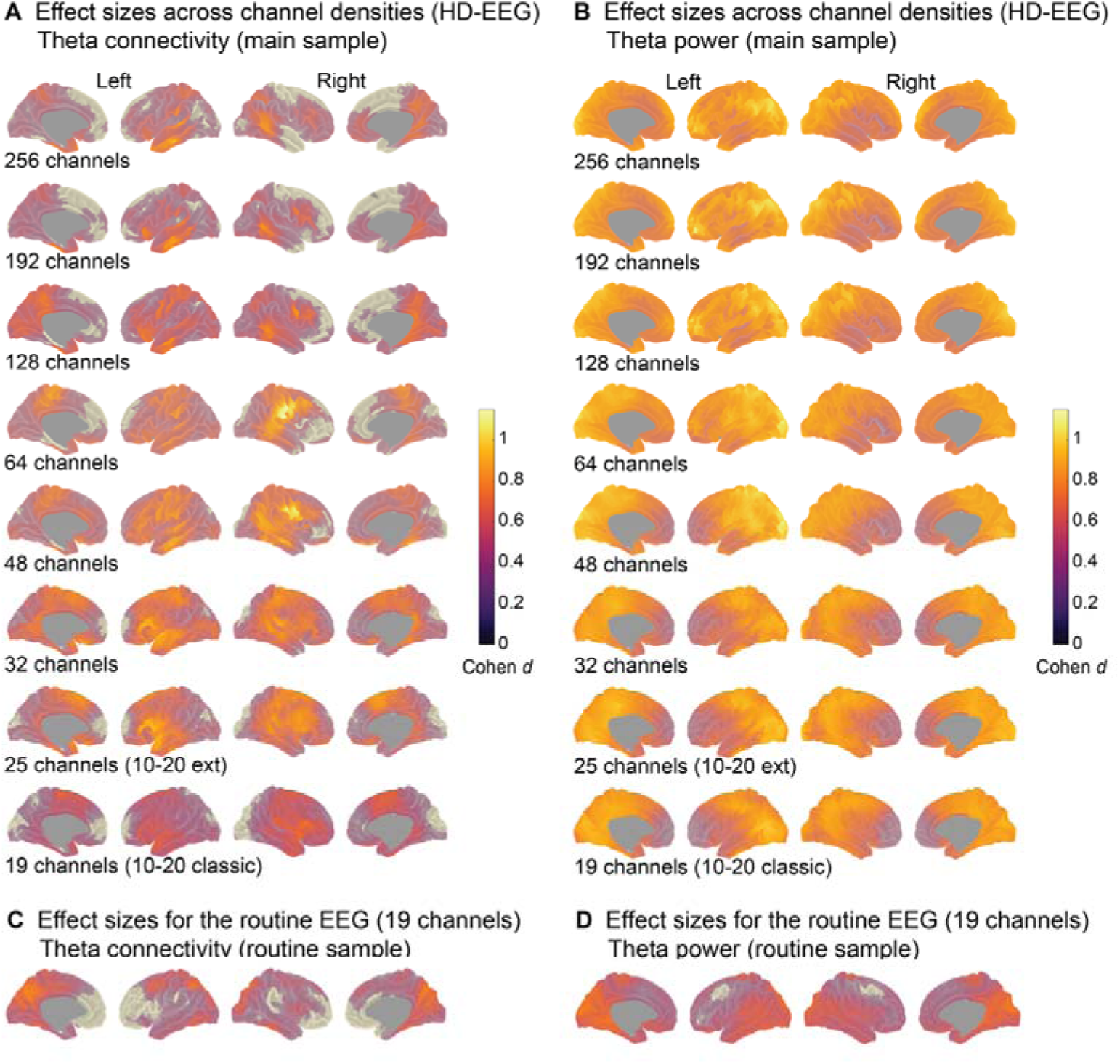
Cohen d maps of vertex-based group comparisons (IGE > controls) for the HD-EEG study sample and the routine sample (canonical head models) Shown are standardized effect sizes for increased EEG levels in the patients with IGE compared with the controls. Note, only results for the theta frequency bands are presented and based on source-projected signals using canonical head models in the HD-EEG study sample (A and B) and the routine sample (B and C). All d-values were computed based on the respective t-values of the group factor of the linear models and corrected for the influence of age, sex, and measurement site. d ≥.8 indicates a large, d =.5 a medium, and d =.2 a small effect (Cohen, 1992). Sample sizes HD-EEG: n_IGE_ = 35; n_controls_ = 54; Routine EEG n_IGE_ = 71; n_controls_ = 43.

## 4. Discussion

This study provides a comprehensive overview of global and spatial differences in brain activity of individuals with IGE compared with controls depending on the number of EEG channels, type of head model, and parcellation scheme. We systematically evaluated group-level outcomes and found that channel density had a stronger effect on EEG connectivity than on power, largely independent of the parcellation. The results for the head model type (individual vs. canonical) were similar when conducted at brain vertices, but better concordance was achieved with a regional or network parcellation. In essence, the main electrophysiological signature of increased theta connectivity and power was similarly observed in the high-density and the clinical low-density setting, as validated in two different study samples.

Not many studies have evaluated the impact of spatial sampling on brain-wide approaches in generalized epilepsies before. Silva Alves and colleagues (Silva Alves et al., 2024) have found local topological network reorganizations using 256-channel HD-EEG that were still observable when using only a 25-electrode montage. This suggests that the most prominent effects in EEG signals of patients with IGE can be preserved with a few channels - in that study for approximately 20 individuals per group and for effects in the delta, theta, and beta bands in large cortical areas. Similarly, our analyses show that global differences between patients and controls, hence, the direction of the effect, can be detected with sparse arrays. Global differences were also the strongest in the delta, theta, and beta bands, with Cohen’s d between ∼0.8 and 0.4 for connectivity across channel sets and for power across the spectrum with effect sizes between ∼1.2 and 0.75. These findings are largely in agreement with previous studies demonstrating markedly increased power in IGE (Clemens et al., 2012, Faiman et al., 2021) and increased connectivity (Elshahabi et al., 2015, Li Hegner et al., 2018, Silva Alves et al., 2024, Stier et al., 2022), with strong effects for the theta band. Moreover, we replicated these findings to some extent in a second sample of about 71 patients and 43 controls recorded in a clinical setting using the same methods and a routine, low-density EEG.

However, when it comes to the spatial topography of the found effects, a clear impact of the channel densities was observed. This was mainly the case for the connectivity estimates based on fewer than ∼64 channels. For signal power, reduced spatial coverage was less problematic, still yielding correlation coefficients of ∼0.3 for low-density maps (32-19 channels) with the original HD-EEG set. Crucially, while EEG power is less sensitive to noise and is usually estimated for a single location in the brain, the connectivity metric applied here reflects the overall synchronization between pair-wise signals across the brain. Hence, estimating complex markers such as connectivity is presumably more prone to spatial imprecisions. This aligns with earlier recommendations to use at least 64 channels when graph theoretical metrics are computed (Hatlestad-Hall et al., 2023), which usually involves describing more long-distance network connections. Also, in the context of source localization in focal epilepsies, earlier studies suggested that at least 64 electrodes should be used to minimize the localization error (Song et al., 2015). Here, spatial sensitivity is crucial, whereas, in the search for biomarkers using electrophysiology at the group level, large-scale network alterations are reported with high individual variability and spatial variations across IGE subtypes (Li Hegner et al., 2018, Stier et al., 2022). Indeed, such brain-wide alterations were detected for most of the virtually reduced channel sets here, particularly in temporal and frontocentral areas, but with various effect sizes and more blurred in low-density maps.

Regarding our routine sample, the spatial topography for both the virtual and real 19-channel settings varied to some degree. Also, no group differences were observed for the delta frequency band. Several factors might have caused these differences: First, proportionally more patients with absence seizures (CAE, JAE) were included in the HD-EEG sample than in the clinical sample, and second, more GSWD occurred. Data trials containing GSWD (±10 seconds of data) were excluded for each individual, but it is conceivable that brain activity beyond this period is altered if GSWD are present (Stier et al., 2021, Tangwiriyasakul et al., 2018). Further investigations with larger sample sizes are needed to disentangle network changes across IGE subtypes and other epilepsy syndromes and should address how group-level estimates can inform patient-tailored diagnostics in regular medical care. Third, the routine sample underwent a slightly different recording procedure than the HD-EEG sample (see Methods). Berger maneuvers and hyperventilation might have induced variation in the resting-state despite removing the respective data segments for analysis. Yet, our results suggest that data acquired in clinical settings with low-density and often less costly EEG systems can be informative and should be leveraged for a better understanding of network changes among epilepsies, but spatial precision should be regarded with caution.

Our work further contributes with an evaluation of the head model type and anatomical resolution. A remapping of EEG metrics according to regions or functional networks did not substantially yield more stable results for channel reductions. The only exception was the results for HD-EEG power in combination with a canonical head model. This can partially be explained given that a canonical head model is built on a template brain and not on individually defined tissue boundaries, leading to less precise source estimation (Henson et al., 2009, Huiskamp et al., 1999). Averaging vertex-level data smooths local inaccuracies and leverages group effects. In a similar vein, statistical outcomes tended to be more similar between the head model types with coarser anatomical resolutions. Of note, we used boundary element methods in combination with a simplified three-layer head model. It is conceivable that other methods considering more complex brain geometries (Vorwerk et al., 2018) would generate larger differences between individual and canonical head models.

Finally, it needs to be said that we defined the original 256-channel montage as the gold standard for our analyses. There was a slight tendency of intermediate arrays (∼64-48 channels) resulting in stronger group differences. Theoretically, more channels should improve the analysis, but only if the signal-to-noise ratio is low (da Silva, 2013). In other words, using high-density arrays in presence of a high noise level may blur the underlying true signal. At the same time, more channels yield a higher spatial overlap of leadfields for the source projection, which can lead to ill-defined covariance matrices and needs to be regulated. We kept a regularization of 5% for all channel densities, which should minimize this problem. Overall, the ground truth or the true network alterations in IGE are not known and only indirectly accessible, such that the specificity and validity of interictal resting-state markers require further investigations. This clearly differs from work on spatial sampling in the presurgical setting, for which the success of the diagnostic accuracy and intervention usually is followed up or can be modeled. Nevertheless, as noted above, this and previous studies consistently point to wide-spread alterations in the theta frequency range. Intriguingly, Stevelink and colleagues found genetic correlations for EEG levels in theta and genetic risk for IGE (Stevelink et al., 2021). Others suggest that 5–9 Hz oscillations represent periods of increased seizure susceptibility based on genetic rat models (Nikalexi et al., 2025), further supporting a mechanistic role of theta-band synchrony in the maintenance or propagation of pathological network states in humans. Ji et al. (2025) demonstrated remarkable seizure frequency reduction in patients with IGE upon deep brain stimulation of the centromedian nucleus of the thalamus, which has been identified as a key node in an IGE network.

## 5. Conclusion

To conclude, main group-level effects in individuals with IGE can be reliably estimated using low-density EEG arrays and amplitude-based measures such as power. However, caution is needed when probing the spatial profile of connectivity estimates, as low-density EEG (<64 channels) captures global alterations well but lacks the spatial resolution required for detailed local or network-level analyses. While incorporating individual anatomical data is ideal, canonical head models can serve as a reasonable alternative, albeit with reduced spatial precision.

## Data availability statement

All relevant study results are available from the corresponding author upon request. Raw imaging and electrophysiological data are not publicly available due to data protection regulations. Code used for preprocessing of the EEG data has been deposited in https://github.com/nmri-nfocke/nmri-meg-eeg-pipeline. Analysis code can be accessed via https://github.com/chstier/ChannelDensIGE.

## Acknowledgements

We would like to express our gratitude to all study participants.

## Supporting Information to

**Figure S1.**
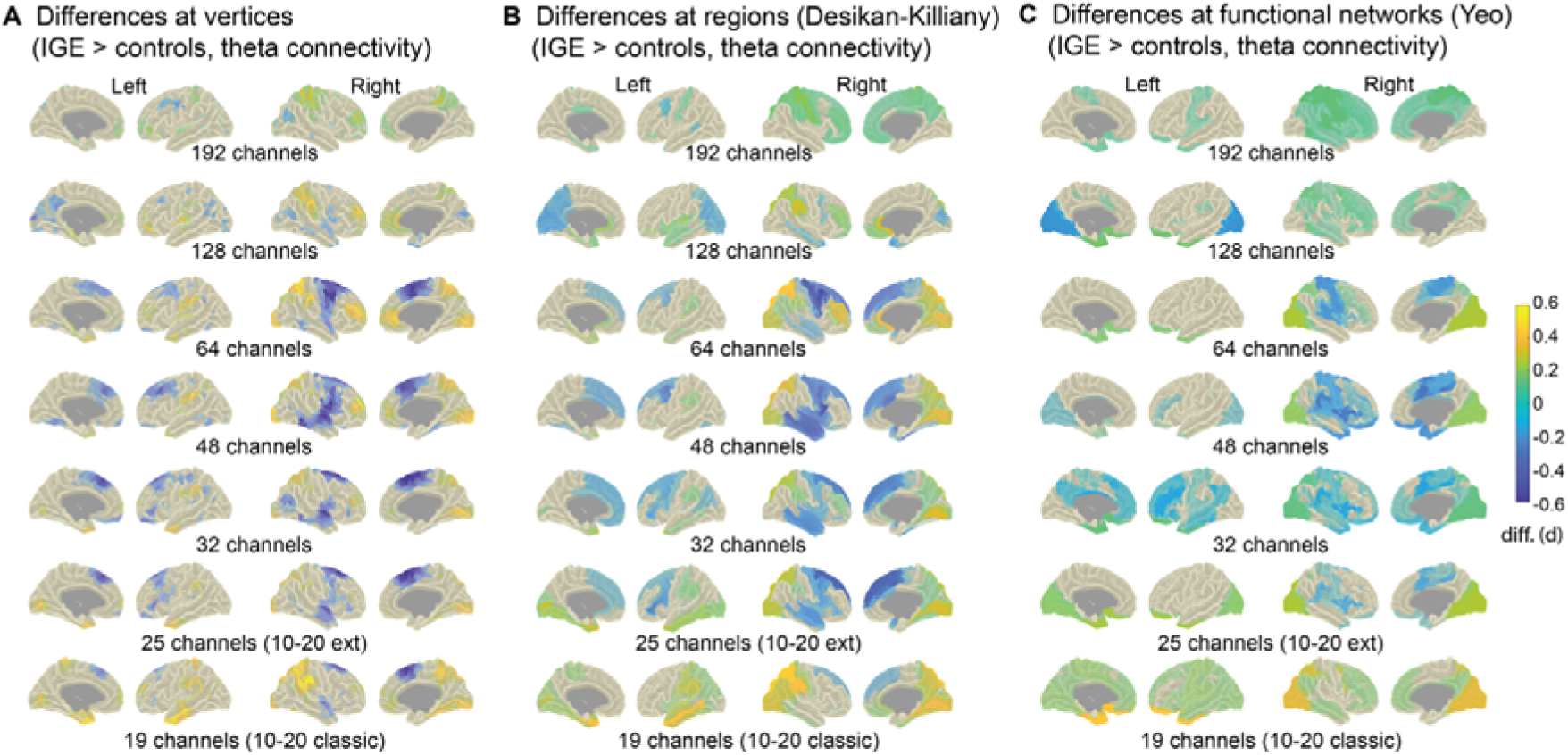
Deviations from the original effect size maps (256 channels, individual head models) for connectivity at different channel sets and parcellations Statistical group comparisons (IGE > controls) were performed separately for different parcellations of the brain (vertices, anatomical regions, and functional networks) based on permutation analysis of linear models, including age, sex, and scanner site as covariates (n_IGE_ = 35; n_controls_ = 54). Standardized effect sizes (Cohen d) were calculated based on the resulting t-values corrected for the influence of the covariates. For each resolution, the differences between the Cohen d-maps for the 256 channel-layout and those for the reduced layouts (19 to 192 channel densities) were then calculated and color-coded (A-C). Green and yellow colors indicate larger effect sizes and blue colors depict smaller effect sizes in the 256-reference map than in the respective lower sampling map. The results shown in (A) rely on EEG signals projected to 2004 vertices using individual head models and different channel compositions. For the anatomical (B) and network-based (C) analyses, individual connectivity values at the vertices were averaged for each region of interest (Desikan et al., 2006; Yeo et al., 2011). diff.(d) = differences in Cohen’s d value.

**Figure S2.**
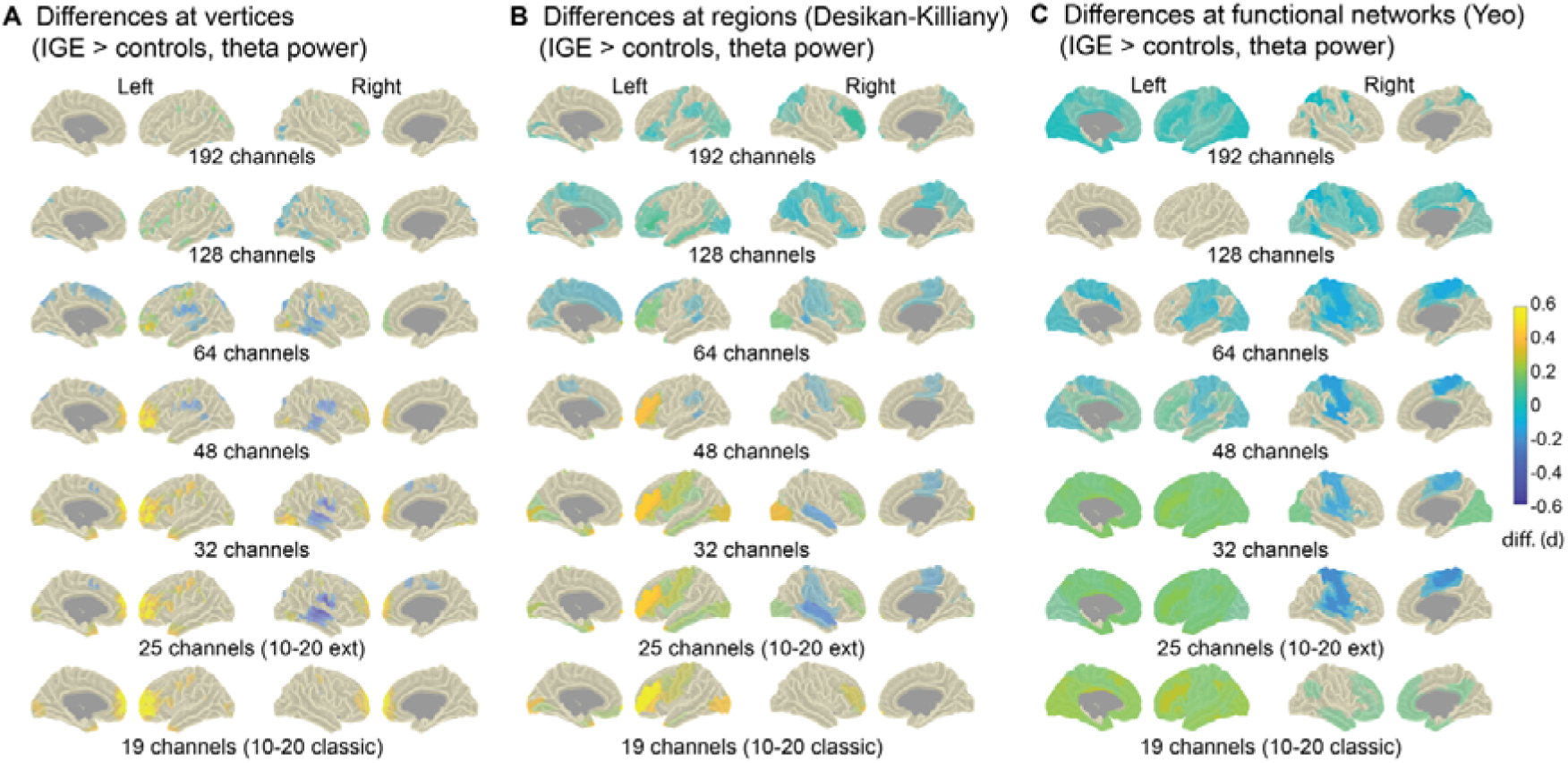
Deviations from the original effect size maps (256 channels, individual head models) for power at different channel sets and parcellations Statistical group comparisons (IGE > controls) were performed separately for different parcellations of the brain (vertices, anatomical regions, and functional networks) based on permutation analysis of linear models, including age, sex, and scanner site as covariates (n_IGE_ = 35; n_controls_ = 54). Standardized effect sizes (Cohen d) were calculated based on the resulting t-values corrected for the influence of the covariates. For each resolution, the differences between the Cohen d-maps for the 256 channel-layout and those for the reduced layouts (19 to 192 channel densities) were then calculated and color-coded (A-C). Green and yellow colors indicate larger effect sizes and blue colors depict smaller effect sizes in the 256-reference map than in the respective lower sampling map. The results shown in (A) rely on EEG signals projected to 2004 vertices using individual head models and different channel compositions. For the anatomical (B) and network-based (C) analyses, individual power values at the vertices were averaged for each region of interest (Desikan et al., 2006; Yeo et al., 2011). diff.(d) = differences in Cohen’s d value.

## References

Alexander-Bloch AF, Shou H, Liu S, Satterthwaite TD, Glahn DC, Shinohara RT, et al. On testing for spatial correspondence between maps of human brain structure and function. Neuroimage 2018;178:540–51.

Ashburner J, Friston KJ. Unified segmentation. Neuroimage 2005;26(3):839–51.

Brodbeck V, Spinelli L, Lascano AM, Wissmeier M, Vargas M-I, Vulliemoz S, et al. Electroencephalographic source imaging: a prospective study of 152 operated epileptic patients. Brain 2011;134(10):2887–97.

Clemens B, Puskás S, Besenyei M, Emri M, Opposits G, Kis S, et al. EEG-LORETA endophenotypes of the common idiopathic generalized epilepsy syndromes. Epilepsy research 2012;99(3):281–92.

Clemens B, Szigeti G, Barta Z. EEG frequency profiles of idiopathic generalised epilepsy syndromes. Epilepsy research 2000;42(2-3):105–15.

Cohen J. A power primer. Psychological bulletin 1992;112(1):155.

da Silva FL. EEG and MEG: relevance to neuroscience. Neuron 2013;80(5):1112–28.

Desikan RS, Ségonne F, Fischl B, Quinn BT, Dickerson BC, Blacker D, et al. An automated labeling system for subdividing the human cerebral cortex on MRI scans into gyral based regions of interest. Neuroimage 2006;31(3):968–80.

Elshahabi A, Klamer S, Sahib AK, Lerche H, Braun C, Focke NK. Magnetoencephalography reveals a widespread increase in network connectivity in idiopathic/genetic generalized epilepsy. PLoS One 2015;10(9):e0138119.

Faiman I, Smith S, Hodsoll J, Young AH, Shotbolt P. Resting-state EEG for the diagnosis of idiopathic epilepsy and psychogenic nonepileptic seizures: A systematic review. Epilepsy & Behavior 2021;121:108047.

Fisher RA. 014: On the“Probable Error” of a Coefficient of Correlation Deduced from a Small Sample. 1921.

Gramfort A, Papadopoulo T, Olivi E, Clerc M. OpenMEEG: opensource software for quasistatic bioelectromagnetics. Biomedical engineering online 2010;9(1):1–20.

Gross J, Kujala J, Hämäläinen M, Timmermann L, Schnitzler A, Salmelin R. Dynamic imaging of coherent sources: studying neural interactions in the human brain. Proceedings of the National Academy of Sciences 2001;98(2):694–9.

Hatlestad-Hall C, Bruña R, Liljeström M, Renvall H, Heuser K, Taubøll E, et al. Reliable evaluation of functional connectivity and graph theory measures in source-level EEG: How many electrodes are enough? Clinical Neurophysiology 2023;150:1–16.

Henson RN, Mattout J, Phillips C, Friston KJ. Selecting forward models for MEG source-reconstruction using model-evidence. Neuroimage 2009;46(1):168–76.

Huiskamp G, Vroeijenstijn M, van Dijk R, Wieneke G, van Huffelen AC. The need for correct realistic geometry in the inverse EEG problem. IEEE Transactions on Biomedical Engineering 1999;46(11):1281–7.

Jasper HH. The 10-20 electrode system of the International Federation. Electroencephalogr Clin Neurophysiol 1958;10:371–5.

Ji G-J, Fox MD, Morton-Dutton M, Wang Y, Sun J, Hu P, et al. A generalized epilepsy network derived from brain abnormalities and deep brain stimulation. Nature communications 2025;16(1):2783.

Justesen AB, Foged MT, Fabricius M, Skaarup C, Hamrouni N, Martens T, et al. Diagnostic yield of high-density versus low-density EEG: The effect of spatial sampling, timing and duration of recording. Clinical Neurophysiology 2019;130(11):2060–4.

Klem GH. The ten-twenty electrode system of the international federation. the internanional federation of clinical nenrophysiology. Electroencephalogr Clin Neurophysiol Suppl 1999;52:3–6.

Kreilkamp BA, Martin P, Bender B, la Fougère C, van de Velden D, Stier C, et al. Big Field of View MRI T1w and FLAIR Template-NMRI225. Scientific Data 2023;10(1):211.

Lantz G, De Peralta RG, Spinelli L, Seeck M, Michel C. Epileptic source localization with high density EEG: how many electrodes are needed? Clinical neurophysiology 2003;114(1):63–9.

Li Hegner Y, Marquetand J, Elshahabi A, Klamer S, Lerche H, Braun C, et al. Increased functional MEG connectivity as a hallmark of MRI-negative focal and generalized epilepsy. Brain topography 2018;31(5):863–74.

Marquetand J, Vannoni S, Carboni M, Li Hegner Y, Stier C, Braun C, et al. Reliability of magnetoencephalography and high-density electroencephalography resting-state functional connectivity metrics. Brain connectivity 2019;9(7):539–53.

Miyauchi T, Endo K, Yamaguchi T, Hagimoto H. Computerized analysis of EEG background activity in epileptic patients. Epilepsia 1991;32(6):870–81.

Nikalexi E, Maksimenko V, Seidenbecher T, Budde T, Pape H-C, Lüttjohann A. Spectral and coupling characteristics of somatosensory cortex and centromedian thalamus differentiate between pre-and inter-ictal 5–9 Hz oscillations in a genetic rat model of absence epilepsy. Neurobiology of Disease 2025;205:106777.

Niso G, Carrasco S, Gudín M, Maestú F, Del-Pozo F, Pereda E. What graph theory actually tells us about resting state interictal MEG epileptic activity. NeuroImage: clinical 2015;8:503–15.

Nolte G, Bai O, Wheaton L, Mari Z, Vorbach S, Hallett M. Identifying true brain interaction from EEG data using the imaginary part of coherency. Clinical neurophysiology 2004;115(10):2292–307.

Oostenveld R, Fries P, Maris E, Schoffelen J-M. FieldTrip: open source software for advanced analysis of MEG, EEG, and invasive electrophysiological data. Computational intelligence and neuroscience 2011;2011.

Pegg EJ, Taylor JR, Mohanraj R. Spectral power of interictal EEG in the diagnosis and prognosis of idiopathic generalized epilepsies. Epilepsy & Behavior 2020;112:107427.

Saad ZS, Reynolds RC. Suma. Neuroimage 2012;62(2):768–73.

Scheffer IE, Berkovic S, Capovilla G, Connolly MB, French J, Guilhoto L, et al. ILAE classification of the epilepsies: position paper of the ILAE Commission for Classification and Terminology. Epilepsia 2017;58(4):512–21.

Seeck M, Koessler L, Bast T, Leijten F, Michel C, Baumgartner C, et al. The standardized EEG electrode array of the IFCN. Clinical neurophysiology 2017;128(10):2070–7.

Segovia-Oropeza M, Rauf EHU, Heide EC, Focke NK. Quantitative EEG signatures in patients with and without epilepsy development after a first seizure. Epilepsia Open 2025;10(2):427–40.

Silva Alves A, Rigoni I, Mégevand P, Lagarde S, Picard F, Seeck M, et al. High-density electroencephalographic functional networks in genetic generalized epilepsy: Preserved whole-brain topology hides local reorganization. Epilepsia 2024;65(4):961–73.

Smith SM, Nichols TE. Threshold-free cluster enhancement: addressing problems of smoothing, threshold dependence and localisation in cluster inference. Neuroimage 2009;44(1):83–98.

Song J, Davey C, Poulsen C, Luu P, Turovets S, Anderson E, et al. EEG source localization: Sensor density and head surface coverage. Journal of neuroscience methods 2015;256:9–21.

Staljanssens W, Strobbe G, Holen RV, Birot G, Gschwind M, Seeck M, et al. Seizure onset zone localization from ictal high-density EEG in refractory focal epilepsy. Brain topography 2017;30:257–71.

Stevelink R, Luykx JJ, Lin BD, Leu C, Lal D, Smith AW, et al. Shared genetic basis between genetic generalized epilepsy and background electroencephalographic oscillations. Epilepsia 2021;62(7):1518–27.

Stier C, Elshahabi A, Hegner YL, Kotikalapudi R, Marquetand J, Braun C, et al. Heritability of magnetoencephalography phenotypes among patients with genetic generalized epilepsy and their siblings. Neurology 2021;97(2):e166–e77.

Stier C, Loose M, Kotikalapudi R, Elshahabi A, Li Hegner Y, Marquetand J, et al. Combined electrophysiological and morphological phenotypes in patients with genetic generalized epilepsy and their healthy siblings. Epilepsia 2022;63(7):1643–57.

Stoyell SM, Wilmskoetter J, Dobrota M, Chinappen DM, Bonilha L, Mintz M, et al. High density EEG in current clinical practice and opportunities for the future. Journal of clinical neurophysiology: official publication of the American Electroencephalographic Society 2021;38(2):112.

Tangwiriyasakul C, Perani S, Abela E, Carmichael DW, Richardson MP. Sensorimotor network hypersynchrony as an endophenotype in families with genetic generalized epilepsy: A resting-state functional magnetic resonance imaging study. Epilepsia 2019;60(3):e14–e9.

Tangwiriyasakul C, Perani S, Centeno M, Yaakub SN, Abela E, Carmichael DW, et al. Dynamic brain network states in human generalized spike-wave discharges. Brain 2018;141(10):2981–94.

Team RC. R: A language and environment for statistical computing. R Foundation for Statistical Computing, Vienna, Austria. http://wwwR-projectorg/ 2013.

Vorderwülbecke BJ, Baroumand AG, Spinelli L, Seeck M, van Mierlo P, Vulliémoz S. Automated interictal source localisation based on high-density EEG. Seizure 2021;92:244–51.

Vorwerk J, Oostenveld R, Piastra MC, Magyari L, Wolters CH. The FieldTrip-SimBio pipeline for EEG forward solutions. Biomedical engineering online 2018;17:1–17.

Winkler AM, Ridgway GR, Douaud G, Nichols TE, Smith SM. Faster permutation inference in brain imaging. Neuroimage 2016;141:502–16.

Winkler AM, Ridgway GR, Webster MA, Smith SM, Nichols TE. Permutation inference for the general linear model. Neuroimage 2014;92:381–97.

Yeo BT, Krienen FM, Sepulcre J, Sabuncu MR, Lashkari D, Hollinshead M, et al. The organization of the human cerebral cortex estimated by intrinsic functional connectivity. Journal of neurophysiology 2011.

## References

Cohen, J. (1992). A power primer. Psychological bulletin, 112(1), 155.

Desikan, R. S., Ségonne, F., Fischl, B., Quinn, B. T., Dickerson, B. C., Blacker, D., Buckner, R. L., Dale, A. M., Maguire, R. P., & Hyman, B. T. (2006). An automated labeling system for subdividing the human cerebral cortex on MRI scans into gyral based regions of interest. Neuroimage, 31(3), 968–980.

Oostenveld, R., Fries, P., Maris, E., & Schoffelen, J.-M. (2011). FieldTrip: open source software for advanced analysis of MEG, EEG, and invasive electrophysiological data. Computational intelligence and neuroscience, 2011.

Yeo, B. T., Krienen, F. M., Sepulcre, J., Sabuncu, M. R., Lashkari, D., Hollinshead, M., Roffman, J. L., Smoller, J. W., Zöllei, L., & Polimeni, J. R. (2011). The organization of the human cerebral cortex estimated by intrinsic functional connectivity. Journal of neurophysiology.

